# Food Insecurity and Food Practices among Child Care Workers During COVID-19

**DOI:** 10.1101/2025.02.20.25322598

**Authors:** Jaclyn M. Dynia, Abel J. Koury, Randi A. Bates

**Affiliations:** The Ohio State University, Columbus, OH, United States; Compelling Analytics, LLC., Austin, TX, United States; University of Cincinnati, Cincinnati, OH, United States

**Keywords:** Food insecurity, child care workers, early childhood, low-wage workers, COVID-19

## Abstract

During the COVID-19 pandemic, food insecurity skyrocketed among U.S. households. In particular, child care workers may have been severely affected, with 33% reporting food insecurity (RAPID-EC, 2021). Using nationally representative data from December 2020, roughly one year into the COVID-19 pandemic, we conducted a detailed examination of food insecurity and practices among U.S. child care workers. We found that child care workers experienced triple the rates of food insecurity compared to the general population (33.3% versus 10.2%), with 20% reporting high/very high food insecurity. To cope with food insecurity, child care workers were more likely to use individual-level strategies (e.g., stretching food budgets) than participating in local or federal programs. After accounting for demographic characteristics, higher-income households had lower odds of experiencing food insecurity, suggesting that increasing wages for child care workers, especially during periods of economic turmoil, could prevent food insecurity among this vital workforce.

---

Food insecurity, or inadequate access to enough nutritious food to maintain a healthy and active life, directly and indirectly impacts all health domains. Food insecurity is associated with an increased risk of hypertension, diabetes, asthma, and depression (e.g., Gregory & Coleman-Jensen, 2017; Noonan et al., 2016) as well as poor sleep and diet (Gundersen & Ziliak, 2015), which exacerbate chronic health conditions. Currently, 17 million households experience some food insecurity (Coleman-Jensen et al., 2021). Despite a reassuring decline in food insecurity over the last decade, rates rose dramatically in 2020 due to the COVID-19 pandemic and have yet to return to pre-pandemic levels (Coleman-Jensen et al., 2021). However, these rates mask health equity concerns, namely heterogeneity by race and ethnicity, household composition, and employment. Food insecurity is more common among households with those of color, children, and those led by un- or under-employed individuals (Coleman-Jensen et al., 2021). Intersectionality may also exacerbate problems with food insecurity. For example, low-wage workers, especially those from traditionally marginalized racial and ethnic groups, are more likely to experience food insecurity and at more severe levels than those from higher socioeconomic backgrounds (Coleman-Jensen et al., 2021; Morita & Kato, 2023).

One sector of low-wage workers vulnerable to food insecurity are child care workers. Child care workers are an essential workforce and attend to and educate children in various settings, such as childcare institutions and private households (Bureau of Labor Statistics, 2022b). Yet, child care workers earn less than 98% of all occupations (McLean et al., 2021) and earn roughly half the hourly wage of the average U.S. worker ($13.22 vs. $27.31; Banerjee et al., 2021). Even in the highest-paying states, child care workers earn less than $38,000 on average (Bureau of Labor Statistics, 2022a). These wages are generally not considered *livable* (Jiang et al., 2017; McLean et al., 2021), resulting in over half of child care workers using public assistance from 2014-2016, including the Supplemental Nutrition Assistance Program (SNAP).

Child care workers’ devastatingly low wages likely contributed to their high rates of food insecurity before and during the pandemic. Before the pandemic, child care workers reported higher rates of food insecurity and at more severe levels than the general population (Loh et al., 2021; McKelvey et al., 2017; Otten et al., 2019). During the pandemic, one estimate from a national survey showed that child care workers’ rates of food insecurity increased from 25% to 33%; for child care workers living below the poverty line, the food insecurity rate doubled (RAPID-EC, 2021). Some of the increase in food insecurity may have resulted from the closing or reduced operations of child care centers across the U.S., leading to the loss of employment and income of nearly one million child care workers (McLean et al., 2021; National Association for the Education of Young Children, 2021).

## Impacts of Child Care Workers’ Food Insecurity

When child care workers experience food insecurity, the subsequent impacts on their physical and mental health are felt by the children in their care, other caregivers in the workforce, and the entire economy. Food insecurity may compromise a caregiver’s ability to provide warm and responsive interactions, which are essential for promoting positive social, emotional, and cognitive development during early childhood (Center on the Developing Child, 2021). Indeed, parents experiencing food insecurity are more easily frustrated and less responsive to their children’s needs (Ashiabi & O’Neal, 2007); it stands to reason that these findings would extend to child care workers tasked with providing warm and responsive interactions to the children in their care.

Additionally, mental health has long been linked to parenting practices, and child care workers report 2-5x greater depressive symptoms than the national average (Linnan et al., 2017). As such, a child care worker who is experiencing depression *and* food insecurity (the two often co-occur; see Koury et al., 2020) may be especially limited in their ability to provide positive interactions that promote children’s development. The impact of lower-quality care (e.g., harsh, inconsistent, and under-stimulating) cannot be overstated. Children enter child care between birth and age five, children’s most vulnerable and rapid period of brain development, when their brains are most susceptible to environmental inputs, or lack thereof (National Research Council, 2012; Whitebook et al., 2018). Children receiving high-quality child care enter kindergarten on stronger footing than their peers, an advantage that sets them up for continued success throughout their lives (Gray-Lobe et al., 2021).

Regarding the economy, child care workers are essential to the U.S. labor market, allowing caregivers (especially mothers) to work (Schochet, 2019). The pandemic illuminated how critical child care workers are to the economy, especially women’s labor force participation. When school and child care centers closed, millions of families, particularly women, were forced to leave the workforce to care for their children (e.g., Kinsey et al., 2020). The disruption to the child care industry also directly impacted women because it is an industry dominated by women, particularly women of color (Dow et al., 2023).

## Current study

In this study, we probed food insecurity and coping strategies among child care workers during the height of the COVID-19 pandemic, in December 2020. We highlight child care workers’ food insecurity for the reasons detailed above, but primarily because they are among the lowest-paid workers in the U.S., despite the essential role they play for families and the economy.

Our study makes two major contributions to the field. First, we examine food insecurity among child care workers with a nationally representative sample. Most of what we know about child care workers’ food insecurity is from subset of states (e.g., Otten et al., 2019) or national studies with non-representative samples (e.g., RAPID-EC). It is thus unclear whether prior results are the experience of most child care workers or only those included in these studies. Second, we define food insecurity among child care workers when food insecurity rates in the U.S. tripled (Fitzpatrick et al., 2021). That is, we chose to examine food insecurity among child care workers roughly one year into the COVID-19 pandemic, when the already strained child care system experienced amplified stressors, with forced closings and massive drops in attendance (National Association for the Education of Young Children, 2020). By understanding the predictors of food insecurity and coping strategies among essential low-wage workers during an economic downturn, policymakers and public health professionals may be better prepared to mitigate hunger during the next economic decline.

## Method

### Data and Sample

This study uses December 2020 data from the Current Population Survey (CPS), a probability sample of approximately 54,000 households selected to represent the entire civilian population in the U.S. (Coleman-Jensen et al., 2021). Households were interviewed once during four consecutive months in a calendar year and then again during the same four months the following year. These rich surveys collect data on the demographic landscape of U.S. households, including employment and economic indicators of well-being. Our primary indicators of interest (e.g., food insecurity, food practices,) were drawn from the Current Population Survey Food Security Supplement (CPS-FSS), the source of annual federal statistics on household food insecurity. Roughly 34,000 households completed the CPS-FSS in addition to the basic CPS interviews (Coleman-Jensen et al., 2021). Households that did not wish to complete the CPS-FSS could opt out (∼24%). Our analytic sample consists of households where the respondent is a child care worker and is at least 18 years of age. Child care workers were identified using the labor code in the CPS-FSS (“child day care services”; industry code 6244). This is under the “Educational, Health, and Social Services” sector (U.S. Census Bureau, 2020). All analyses were weighted to keep results nationally representative of households in the U.S. in December 2020.

### Measures

#### Household food security

Food security was represented with 18 questions to calculate the 12-month Food Security Scales (Household Food Security Scale, 12-Month Reference Period) and 30-day food security scales (U.S. Census Bureau, 2020). These scales are used for the U.S. Department of Agriculture’s annual food security reports (U.S. Census Bureau, 2020). Food security questions addressed the sufficiency of food including whether those in the home skip meals or reduce food intake because there was not enough money for food and how often these events happened over the last 12 months. The final scale used in our analyses was categorical, classifying households on a scale of 1 (*high food security*) to 4 (*very low food security*). Our regression model used a binary version of the variable above, with those experiencing high food security as “0” and anyone experiencing marginal to very low food security as “1”. For ease of interpretation, we discuss our results of marginal-very low food security as food *insecurity*.

#### Food practices and coping

The CPS included additional items on food sufficiency, expenditures, and ways of coping with food insecurity including food programs and individual-level strategies (e.g., stretching one’s food budget). To understand food practices, we examined where child care workers purchased food and how much they spent on food during the previous week (scored *supermarkets*, *grocery stores, restaurants, convenience stores,* and *others)*. To understand how child care workers coped with food insecurity, we examined whether and how often they tried to stretch their food budget, drew upon community programs or food banks, or utilized federal programs (e.g., SNAP).

#### Control variables

We included a series of covariates that have been related to food security, including marital status (*married* vs. *all else*), race and ethnicity (*White, Black, Hispanic,* and *Other Race/Ethnicity*), educational attainment (*below high school, high school, some college, Bachelors or above*), household income (categorical range from *less than $5,000* to *$150,000 or more*), and number of people in the household. Household income was defined as income from the previous year from all members of the household, including money from jobs, dividends, and social security payments.

### Analysis

Our first analysis documented nationally representative statistics on child care workers’ food insecurity, food practices, and coping with food insecurity. Our next analysis utilized weighted logistic regression to predict child care workers’ odds of experiencing food insecurity, based on characteristics listed above.

#### Analytic weights

All analyses were run in Stata 16 using the ‘syvset’ option to apply the appropriate analytic weights (i.e., HHSUPWGT). We restricted the sample to one record per household (see U.S. Census Bureau, 2020 for more details). The weights provided by the Census Bureau have four implied decimal points; as such, to analyze the data by units (i.e., households), each weight was divided by 10,000.

## Results

### Sample characteristics

Our unweighted sample included 226 households in which respondents identified as child care workers of at least 18 years of age. See Table 1 for weighted descriptive statistics. Roughly 96% of child care workers self-identified as female. Child care workers were, on average, 44.7 years old (*SD* = 14.54); ages ranged from 18 to 80. Child care workers largely self-identified as either a White (78.7%) or Black (16.6%) race. We collapsed the bi- or multi-racial, Asian, American Indian or Alaskan Native, and Hawaiian or Pacific Islander into an “Other race” category for our analyses, given the small sample sizes. Nearly a quarter of child care workers identified as Hispanic (23%). Regarding education, most child care workers had some postsecondary education; 45% attended some college or earned an Associate’s degree and roughly 24% earned a Bachelor’s degree or higher. Roughly 30% of child care workers had a high school diploma or below. For marital status, nearly 40% were currently married, and about equal proportions were either never married or divorced/separated/widowed. Geographically, 42% of child care workers were living in the South, with roughly equal proportions in the Midwest, West, and Northwest.

**Table 1.**
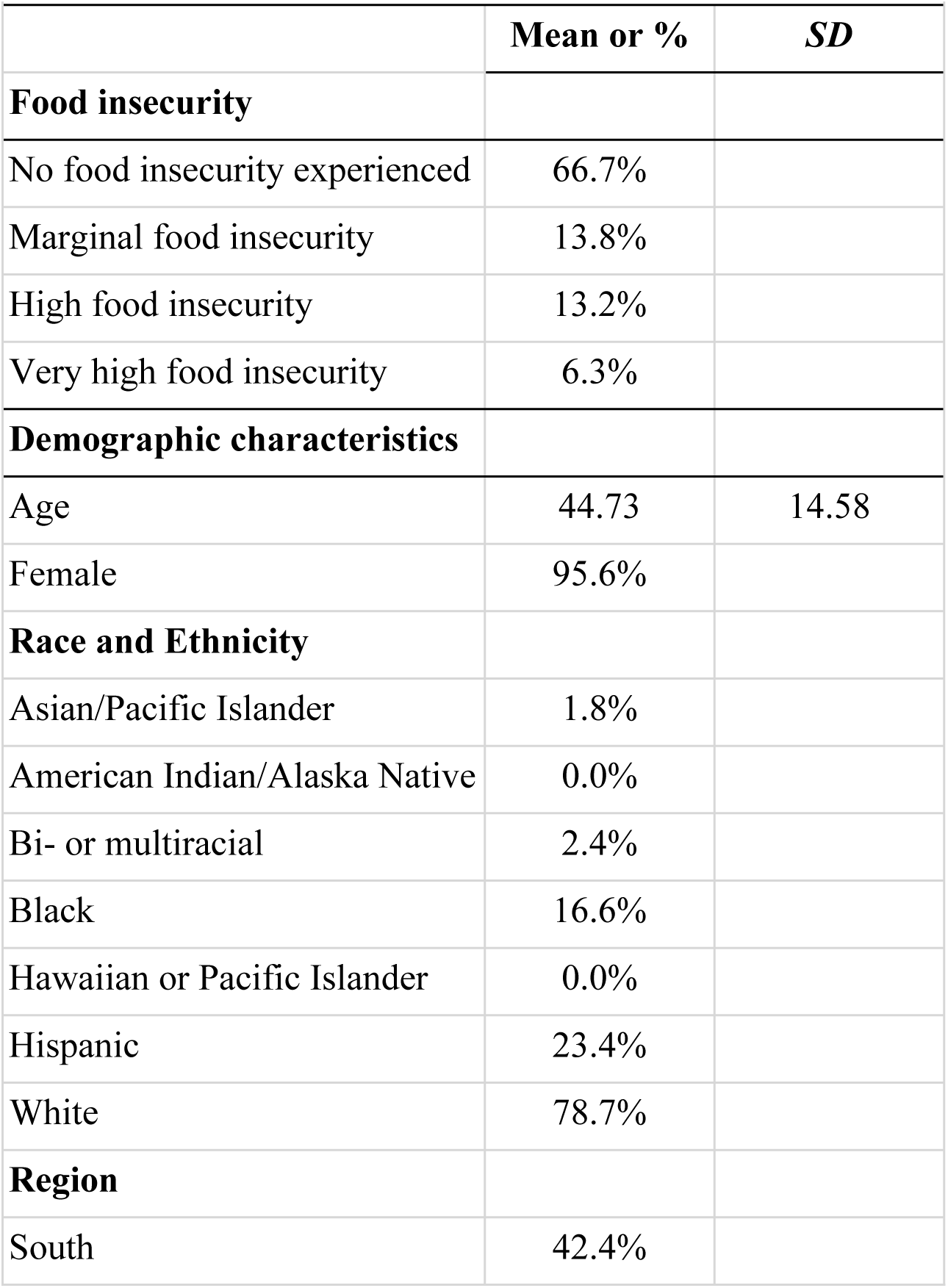
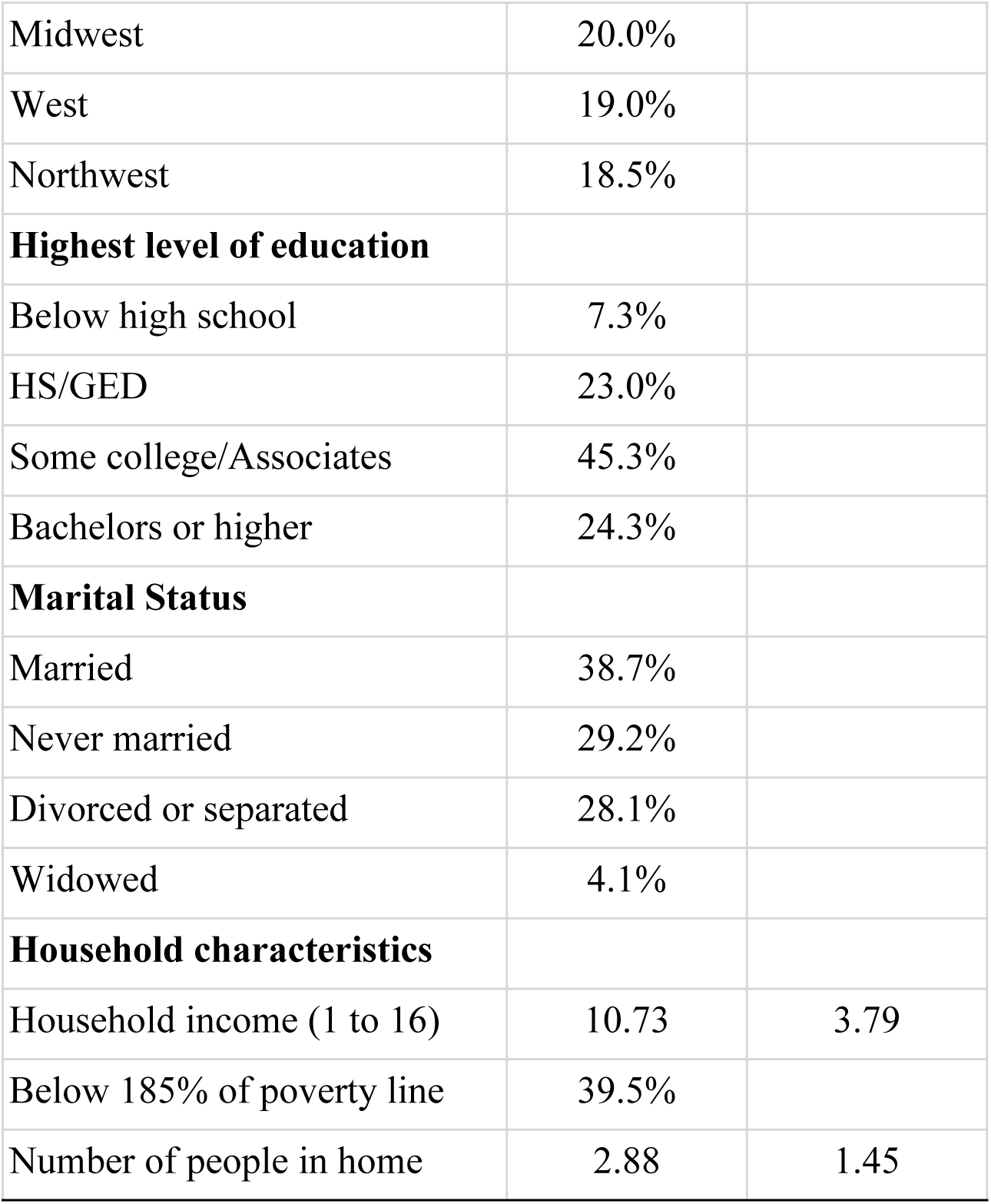
Weighted Descriptive Statistics.

Household size varied widely, from 1-9 individuals (*M* = 2.88, *SD* = 1.45). Most households had between 1 and 4 individuals. Household income ranged widely from less than $5,000 to over $150,000, with an average of $35,000 to $39,999 (median = $40,000 to $49,000). About 25% of child care workers earned less than $25,000, and 22% earned over $75,000. Forty percent of child care workers had a household income below 185% of the federal poverty guidelines, indicating they were experiencing some poverty.

### What are child care workers’ food insecurity and food practices?

First, we examined levels of food insecurity. Notably, 33% of child care workers reported that their household experienced some level of food insecurity over the past year. Overall, 6.3% of individuals reported *very high* levels of food insecurity, 13.2% experienced *high* levels of food insecurity, 13.8% experienced *marginal* food insecurity, and 66.7% did not report any level of food insecurity. Therefore, approximately one in three child care workers experienced some food insecurity and nearly one in five reported high to very high food insecurity.

We examined responses to where food was purchased and how much was spent on food during the previous week, found in Table 2. Over 90% of child care workers bought food in supermarkets or grocery stores. Many child care workers also purchased food at restaurants, fast food places, cafeterias^1^, or vending machines (60.7%). About one-third of child care workers purchased food from other settings such as meat markets, produce stands, bakeries, warehouse clubs, and convenience stores (31.5%). Child care workers spent an average of $157.47 on food during a typical week, with some families spending over twice this amount per week (*S.D.* = $112.00, Range = $0.00 to $450.00). Approximately 11% of child care workers spent $0.00 on food the previous week.

**Table 2.**
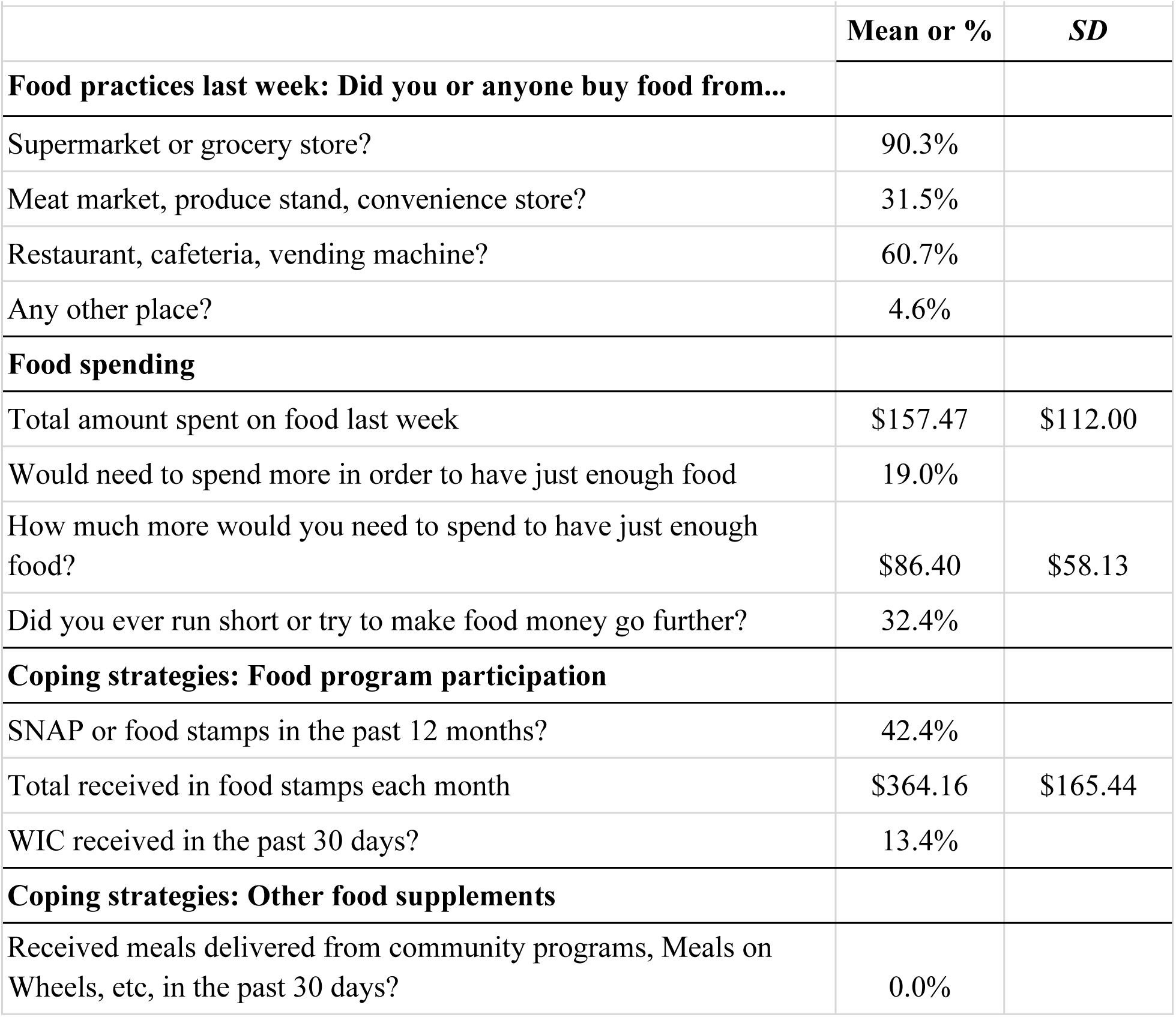
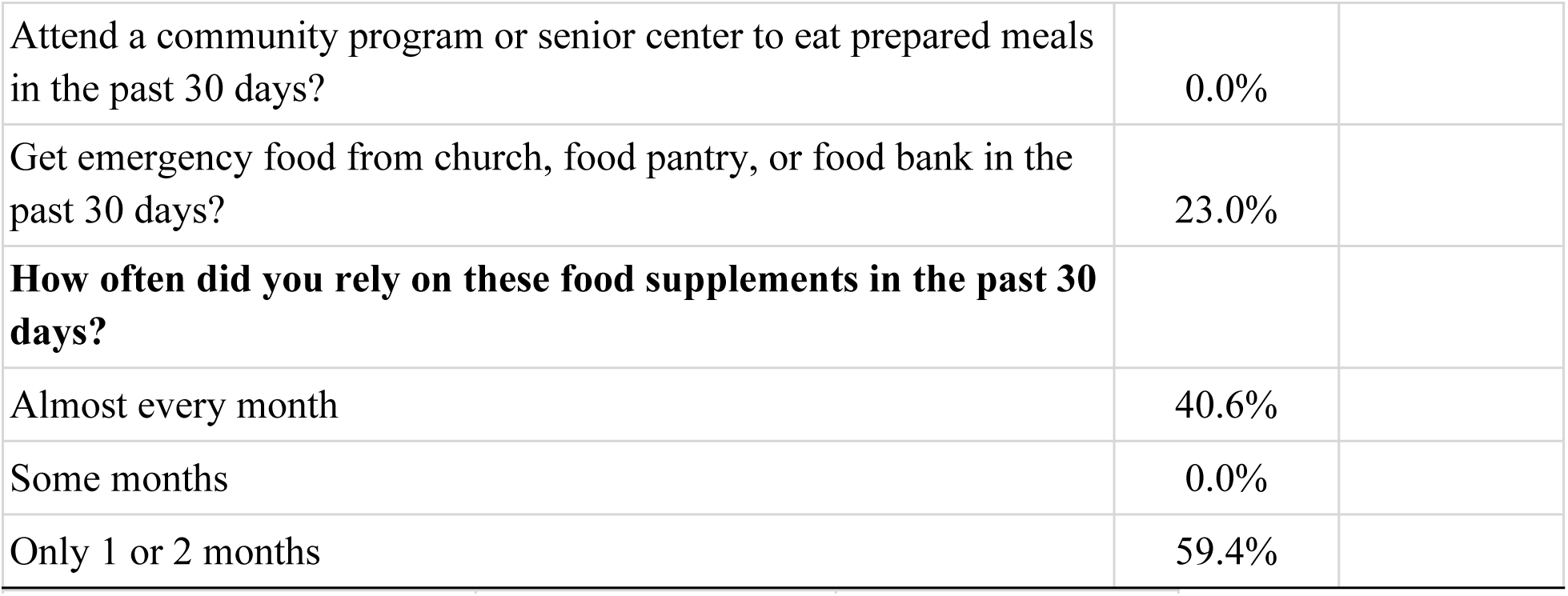
Weighted descriptives: Food practices, spending, and coping strategies.

### Coping with food insecurity

We next examined child care workers’ responses to questions on the minimum amount of money needed for food, ways of coping with food insecurity, and ratings of food insecurity over the previous year. Results are in Table 2.

To stave off food insecurity, results suggest that 19% needed more money each week to purchase food. More specifically, child care workers indicated they would need an average of $86.40 (*S.D.* = $58.13, Range = $20 to $200) each week to buy *just enough* food to meet their household’s needs. Almost one-third of child care workers reported running out of money and trying to stretch their food or food budget in the last year (32.4%).

Despite a sizeable proportion reporting the need for more money for food, only about a third received Supplemental Nutrition Assistance Program (SNAP) or food stamp benefits over the last year. Of the households experiencing the lowest two levels of food security, only 46% received SNAP or food stamp benefits. Those who did use SNAP received an average of an additional $364.16 (*S.D.* = $165.44, range = $20 to $648) in monthly food stamp benefits. A smaller percentage of child care workers reported receiving food through the Women, Infants, and Children (WIC) program (13.4%). The most utilized federal program was school breakfast and lunch. In fact, 90% of children in child care workers’ homes received free or reduced-price breakfast within the past month, and 37.8% received free or reduced-priced lunch. When child care workers needed emergency food, they were most likely to reach out to churches, food banks, and food pantries rather than community programs such as Meals on Wheels. Churches, food banks, and food pantries provided emergency food for roughly a quarter of child care workers, with a sizeable proportion of child care workers utilizing these resources monthly (40%).

### Predictors of Food Insecurity

Using a weighted logistic regression model, we estimated the association between several variables and the binary household food insecurity variable, shown in Table 3. We selected covariates shown to be important predictors of food insecurity in the prior literature and were not collinear with one another (i.e., we included household income, but not “income below the federal poverty line”). The reference category is *high food security*. We found that household income was negatively associated with food insecurity; as income increased by one unit (i.e., the income moved to the next categorical level), the odds of experiencing food insecurity decreased by 16%. The only other significant predictor in the model was for the ethnicity covariate, Hispanic, suggesting that those who reported being Hispanic were over four times as likely to report any level of food insecurity compared to those who are non-Hispanic.

**Table 3.** Logistic Regression of Food Insecurity Among Child Care Workers in December 2020.

|  | **OR** | **SE** |
| --- | --- | --- |
| **Food insecurity** |  |  |
| Household income | 0.84* | 0.06 |
| Below high school | 0.42 | 0.40 |
| Some college/Associates | 0.84 | 0.68 |
| Bachelor's or higher | 0.82 | 0.74 |
| Other | 2.80 | 1.94 |
| Black | 3.52 | 2.53 |
| Hispanic | 4.24** | 2.23 |
| Married | 0.42+ | 0.20 |
| Household size | 0.96 | -0.14 |
| Constant | 2.80 | 3.04 |

*Note.* \* $p < .05$ . \*\* $p < .01$ . + $p < .10$

## Discussion

Food insecurity skyrocketed during the COVID-19 pandemic. Low-wage workers, a group already at heightened risk for food insecurity, experienced much higher food insecurity rates. Our study focused on child care workers, an essential group of low-wage workers who play a critical role in society by educating the future workforce and allowing parents to participate in the labor market. To review, despite the critical nature of their work, child care workers are among the lowest paid workers in the U.S., a fact that likely partially explains their high rates of food insecurity. Our study extended prior research on child care workers’ food insecurity by using a nationally representative dataset and examining predictors of food insecurity during a peak of economic downturn. Our study revealed three main findings: (a) child care workers were more likely to be food insecure during the pandemic than the general population (b) as income increased, the odds of experiencing food insecurity decreased, and (c) Hispanic individuals were four times more likely to be food insecure than non-Hispanic individuals. These findings will be discussed in turn.

### A Third of Child Care Providers Do Not Have Enough Food for Their Households

Using nationally representative data, we found that one-third of child care workers reported experiencing any food insecurity of the past year, with over 19% experiencing the two most severe levels of food insecurity. This rate is nearly double the national average of 10.2% (Coleman-Jenson et al., 2021), highlighting the disproportionate impact of the pandemic on food insecurity for child care workers. Our finding is well-aligned with a convenience survey of over 1,000 child care providers across the U.S. that found child care workers’ food insecurity levels rose from 25% to 33% during the pandemic (RAPID-EC, 2021).

We also found that almost 20% of child care workers expressed the need for additional funds to buy food each week, and another sizable proportion reported running out of money to purchase food at some point during the previous year. Given the high rates of food insecurity and need for additional resources, it is perhaps surprising that only about a third received SNAP or food stamp benefits over the last year. This may be driven by a number of factors including eligibility based on income and household size, but future work could examine whether eligible child care workers are under-utilizing SNAP and other federal programs designed to mitigate food insecurity.

Additionally, child care workers with children reported utilizing free or reduced-price breakfasts and lunches at schools, underscoring the importance of these programs for combating hunger for children in lower-income homes. Food banks, pantries, and churches also helped child care workers access food to feed their families.

### Predictors of Food Insecurity

Results of our logistic regression revealed that household income was a strong predictor of food security. As income increased, the odds of experiencing food insecurity decreased by 16%. Although unsurprising, this finding does highlight that increased wages among child care workers (and other low-wage workers) may prevent food insecurity. This finding aligns with other studies on child care workers and income. For example, RAPID-EC (2022) found that child care workers experiencing poverty also experienced higher levels of food insecurity. Low wages coupled with food insecurity may be especially damaging to child care workers’ health, creating chronic stress, which results in “wear and tear” on the body, and later chronic disease (Juster et al., 2010). Chronic stress and food insecurity together may limit a child care worker’s ability to provide nurturing, responsive care to children, which compromises children’s early development. Thus, increasing wages for child care workers may promote not only child care workers’ health and wellbeing, but also for children in their care.

Finally, those who reported being Hispanic were over four times as likely to report any level of food insecurity compared to those who were non-Hispanic. This corroborates other work finding higher than average rates of food insecurity for Hispanic households (Gundersen et al., 2011). Hispanic child care workers also experience wage gaps compared to white child care workers (Austin et al., 2019; Lee et al., 2023), which may partially drive the higher rates of food insecurity among these workers. It is important to note that women of color make up a sizable proportion of child care workers, especially in home-based settings (Paschall et al., 2020). As such, more work should be directed to understanding how to eliminate food insecurity among child care workers of color.

### Limitations and Future Research

Although this study has provided critical findings on the food insecurity of child care workers using a nationally-representative sample, there are two limitations worth mentioning. First, the sample of child care workers that participated in the CPS was small. Using analytic weights allowed us to derive nationally representative estimates, but future studies are encouraged to oversample child care workers of color to obtain more representative estimates. Second, we identified child care workers by using the labor code in the CPS-FSS (“child day care services”). There is little information regarding who make up those in “child day care services”, including whether these workers are in child care centers, home settings, or public schools. Future studies are encouraged to examine how food insecurity may differ by child care setting.

In sum, child care workers are necessary for a robust economy and provide essential care and education to our youngest learners. Nevertheless, child care workers experience higher rates of food insecurity than the general population, which seems to be related to the low wages and high poverty rates among child care workers. Moreover, experiencing food insecurity can have long-term health consequences. Education policy should continue to focus on workforce development, including increasing child care workers’ wages. We imagine our findings may also extend to understanding other low-wage workers, including how to mitigate food insecurity in these workers.

## Data Availability

The data is US Census data and is publicly available.

## Contributions

Jaclyn M. Dynia: Conceptualization, methodology, writing – original draft, writing – review & editing. Abel J. Koury: Formal analysis, methodology, writing – original draft, writing – review & editing. Randi A. Bates: Writing – original draft, writing – review & editing.

## Funding acknowledgement

During final preparation of the manuscript, Jaclyn M. Dynia’s position was supported by the National Science Foundation Engineering Research Center. Declarations of interest. None.

## Data and code availability

Data is available from the US Census and code is available by the second author.

## Acknowledgements

Jaclyn M. Dynia previous affiliation was SproutFive’s Center for Early Childhood Innovation. Current affiliation is The Ohio State University supported by a National Science Foundation Engineering Research Center.

## Footnotes

1 This includes children in the household who may have bought food from the school cafeteria.

